# Improving Diagnostic Precision: Urine Proteomics Identifies Promising Biomarkers for Necrotizing Enterocolitis

**DOI:** 10.1101/2024.03.21.24304374

**Authors:** Stephen Mackay, Lauren C. Frazer, Grace K. Bailey, Claire M. Miller, Qingqing Gong, Olivia N. DeWitt, Misty Good

## Abstract

**Background:** Necrotizing enterocolitis (NEC) is a severe intestinal disease that primarily impacts preterm infants. Current diagnostic tools are inadequate, so urine proteomics was performed for patients with and without NEC to identify putative biomarkers.

**Research design and methods:** The abundance of urinary proteins detected using an aptamer-based microarray was compared for infants with NEC (*n*=20) and controls, age-matched (*n*=8) or self-matched (*n*=12). Spearman *r* correlation and hierarchical cluster analysis were performed. The area under the curve (AUC) was calculated for receiver operator characteristic curves (ROC).

**Results:** Ninety-nine proteins differed in NEC vs. controls based on median fold change (Log_2_ ± 1.1) and significance (*P* < 0.05). Patterns of abundance were consistent for both types of matching, and samples clustered based on NEC severity. Two panels were built to differentiate between infants with and without NEC. Panel 1 included proteins associated with inflammation/NEC and produced by the intestinal epithelium (REG1B, REG3A, FABP2, DEFA5, AUC 0.90). Panel 2 consisted of proteins with the largest fold change between NEC vs. controls and the highest individual AUC values (REG1B, SSBP1, CRYZL1, ITM2B, IL36B, IL36RN, AUC 0.98).

**Conclusions:** Urine proteins significantly differ between infants with and without NEC, which supports their potential as future biomarkers.

**Graphical abstract.** Overview of study findings. Created with Biorender.com

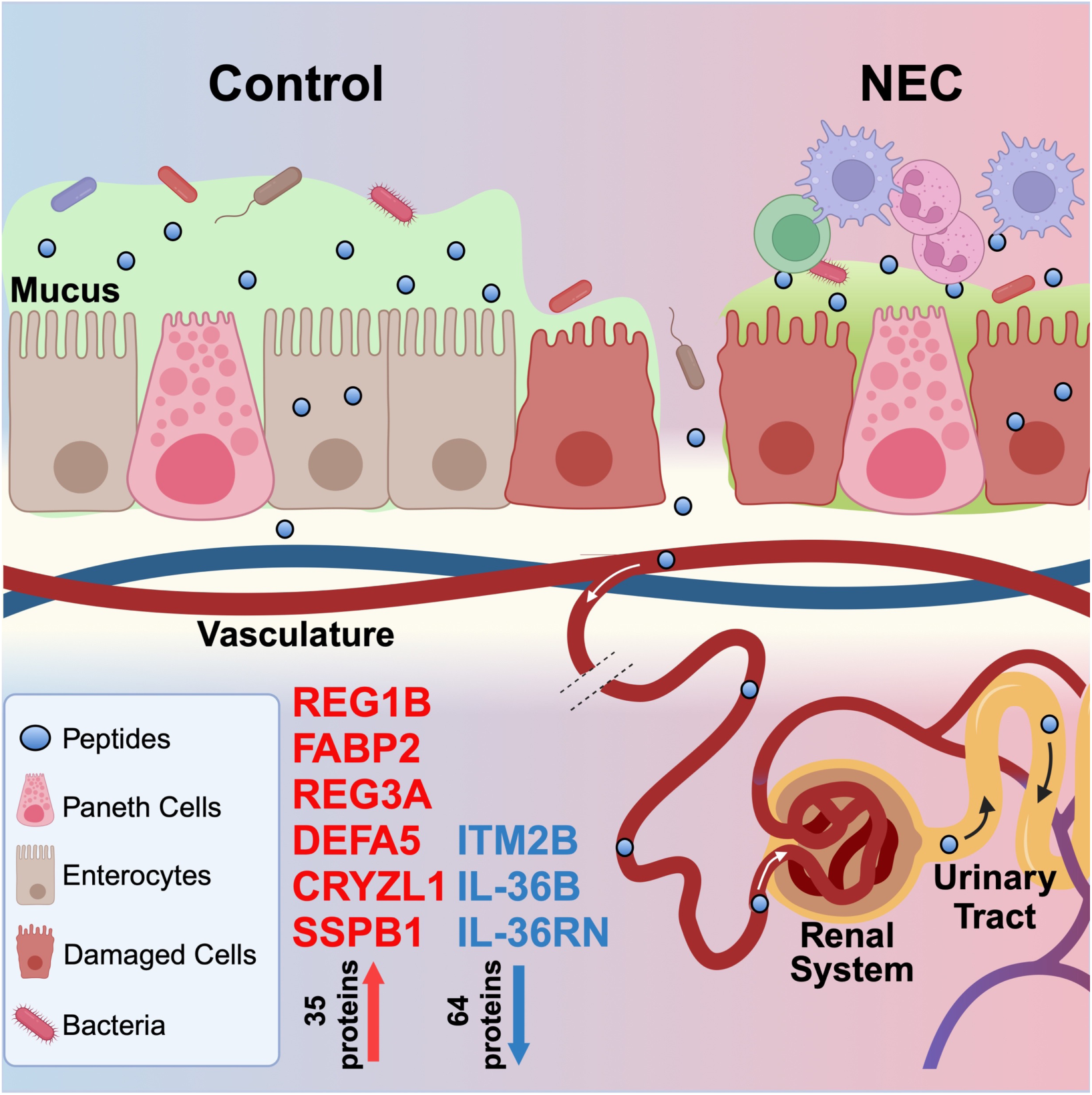

## Background

Necrotizing enterocolitis (NEC) is a rapidly progressive intestinal disease that occurs primarily in preterm and low birth weight neonates and has an incidence of ∼7% for neonates born at < 32 weeks gestation [1]. The first symptoms of NEC, such as feeding intolerance and abdominal distension, can be difficult to distinguish from other etiologies; however, within hours of disease onset, an infant with NEC can progress to needing emergency surgery or death. The rapidity of this clinical decompensation is why an early and accurate diagnosis of NEC is essential. Currently, NEC is diagnosed using imaging modalities such as abdominal X-rays and/or ultrasound combined with physical exams and laboratory tests, but these tools have inadequate sensitivity and specificity, particularly early in the disease course [2]. Unfortunately, no biomarkers for NEC have been implemented in routine clinical practice, which could improve upon some of the current diagnostic limitations.

Identification of biomarkers for NEC has been hindered by a multitude of factors, with the primary barrier being an incomplete understanding of disease pathogenesis. NEC is thought to result from a poorly controlled immune response in the neonatal intestine in the setting of a dysbiotic microbiome. This inflammation leads to intestinal epithelial injury, loss of mucosal barrier integrity, and, ultimately, tissue necrosis [3–6]. The precise factors that induce and exacerbate disease remain an area of active investigation, and that knowledge barrier continues to impede biomarker development.

Several potential biomarkers have been identified for NEC, including proteins/peptides [7,8], metabolites [9], and non-coding RNAs [10,11], but reproducibility and validation of the biomarkers in large cohorts have not been conclusive. In addition, some urinary biomarkers for NEC have been identified, such as fatty acid binding protein-2 (FABP2) [12]; however, these markers are not in use clinically, as normalization of urine samples has been problematic [12]. Optimally, a biomarker for NEC would identify infants early in the disease process and reliably differentiate between neonates with NEC and other intestinal or systemic inflammatory pathologies. In addition, an ideal biomarker would be validated for use with biological specimens that are readily available and non-invasively acquired, given the difficulties associated with acquiring specimens from medically fragile preterm neonates. To overcome the difficulty in obtaining a large number of infants with NEC at one center, we established the multi-center NEC Biorepository to identify biomarkers of disease [13,14].

With improvements in technology, the identification of new biomarkers for NEC is now a distinct possibility. Traditional methods for biomarker identification include quantitative and qualitative analysis of proteins and metabolites using untargeted gas or liquid chromatography coupled with tandem mass spectrometry (LC-MS/MS). Recently, aptamer-based proteomic screens using large, highly selective libraries have been utilized to screen samples for biomarkers in various tissue and biological specimens, including serum, stool, and urine [15–17]. Compared to LC-MS/MS, aptamer-based scans are better able to detect small proteins [18], which presents the opportunity to detect new biomarkers for NEC.

In this study, we utilized aptamer-based proteomics technology (SomaScan^®^) to measure the relative abundance of over 7000 proteins in the urine of infants with NEC and controls. We identified ninety-nine proteins that differed between the groups and performed a detailed analysis of protein abundance, patterns across samples, disease severity, and diagnostic utility. In addition, we identified two protein panels and a pair of proteins that effectively differentiate between infants with NEC and controls. This study provides the foundation for future work investigating urine biomarkers for NEC in a larger population of preterm infants and provides an advance toward identifying urgently needed biomarkers for this devastating disease.

## Methods

### Study design

Samples were prospectively collected from infants admitted to the St. Louis Children’s Hospital Neonatal Intensive Care Unit (NICU) in St. Louis, Missouri, USA. Enrollment of infants 22 to 42 weeks gestation was conducted according to protocols approved by the Washington University in St. Louis School of Medicine Institutional Review Board (IRB protocol numbers 201706182 and 201802101). Infants with major congenital anomalies were excluded. For the current study, we included infants initially enrolled as controls and then subsequently developed NEC (self-matched, *n* = 12). We also analyzed a cohort of age-matched infants (*n* = 8 pairs), which included infants enrolled at the time of NEC diagnosis (*n* = 8) and their age-matched controls (*n* = 8). Infants with urinary tract infections were excluded. The cohort was comprised of infants born between 24 and 36 weeks gestation (*n* = 28) (**Table 1**) and modified Bell’s staging was used to categorize NEC severity [19–21].

**Table 1.** Study cohort characteristics

| Sample ID |  | Pregnancy and delivery details |  |  |  | Disease severity |  | Surgical intervention |  |  |
| --- | --- | --- | --- | --- | --- | --- | --- | --- | --- | --- |
| Sample ID | NEC/ Control | Gestational age range (weeks) | Biological sex | Birth weight (g) | Route of delivery | NEC | Highest Bell's stage | Radiologic Findings | Surgical NEC | Final disposition |
| **Age-Matched** |  |  |  |  |  |  |  |  |  |  |
| AM 1 | N | 23 - 27 | Male | 480 | CS | Yes | IIB | PN |  | Death, LD |
| AM 1 | C | 33 - 37 | Female | 1940 | V | No |  |  |  | Discharged |
| AM 2 | N | 33 - 37 | Female | 2360 | CS | Yes | IIA | PN |  | Discharged |
| AM 2 | C | 33 - 37 | Female | 1740 | CS | No |  |  |  | Discharged |
| AM 3 | N | 33 - 37 | Female | 2330 | CS | Yes | IIB | PN, PVG |  | Discharged |
| AM 3 | C | 33 - 37 | Male | 2360 | CS | No |  |  |  | Discharged |
| AM 4 | N | 23 - 27 | Male | 640 | CS | Yes | IIB | PN |  | Discharged |
| AM 4 | C | 23 - 27 | Female | 710 | CS | No |  |  |  | Discharged |
| AM 5 | N | 28 - 32 | Female | 1190 | CS | Yes | IIB | PN, PVG |  | Discharged |
| AM 5 | C | 33 - 37 | Male | 2030 | V | No |  |  |  | Discharged |
| AM 6 | N | 23 - 27 | Male | 700 | CS | Yes | IIIB | G | PR, PD | Death, LD |
| AM 6 | C | 23 - 27 | Male | 1150 | V | No |  |  |  | Discharged |
| AM 7 | N | 28 - 32 | Female | 1870 | CS | Yes | IIA | PN |  | Discharged |
| AM 7 | C | 28 - 32 | Female | 2320 | V | No |  |  |  | Discharged |
| AM 8 | N | 23 - 27 | Female | 790 | V | Yes | IIB | PN |  | Discharged |
| AM 8 | C | 28 - 32 | Male | 1580 | V | No |  |  |  | Discharged |
| **Self-Matched** |  |  |  |  |  |  |  |  |  |  |
| SM 9 | C+N | 28 - 32 | Male | 1220 | CS | Yes | IIIA | PN, PVG |  | Discharged |
| SM10 | C+N | 23 - 27 | Female | 760 | V | Yes | IIIB | PN, PVG, P | PD | Death, NEC |
| SM11 | C+N | 23 - 27 | Male | 760 | CS | Yes | IIA | PN |  | Discharged |
| SM12 | C+N | 23 - 27 | Male | 790 | CS | Yes | IIIB | P | PR | Discharged |
| SM13 | C+N | 28 - 32 | Female | 1775 | V | Yes | IB |  |  | Discharged |
| SM14 | C+N | 28 - 32 | Male | 1780 | CS | Yes | IIB | PN |  | Discharged |
| SM15 | C+N | 23 - 27 | Female | 690 | CS | Yes | II |  |  | Study closed |
| SM16 | C+N | 23 - 27 | Male | 430 | CS | Yes | IIB | PN |  | Death, LD |
| SM17 | C+N | 28 - 32 | Male | 1770 | CS | Yes | IIB | PN |  | Discharged |
| SM18 | C+N | 23 - 27 | Female | 710 | CS | Yes | IIA | PN |  | Discharged |
| SM19 | C+N | 23 - 27 | Male | 530 | CS | Yes | IIIA | G |  | Death, LD |
| SM20 | C+N | 23 - 27 | Male | 660 | CS | Yes | IIIB | PVG | PR | Study closed |

Delivery route: CS = cesarean Section, V = vaginal; Surgical NEC: PR = partial resection, PD = peritoneal drain; Disposition: LD = lung disease, Radiological findings: PN = pneumatosis, PVG = portal venous gas, P = pneumoperitoneum, G = gasless abdomen, Study closed = PI relocation.

### Sample collection

Urine samples were collected by placing sterile gauze in a clean diaper. Urine-saturated gauze was then collected at a routine care time, placed in a 10 mL syringe, and stored at 4°C until processed by laboratory staff 1-2 times per day. Urine was subsequently extracted from the gauze by squeezing the syringe, aliquoted into cryotubes, and stored at -80°C until analysis. Samples with visible stool contamination were discarded.

### Proteomics assays

#### Sample preparation for SomaScan^®^ and ELISA

Frozen urine samples (−80°C) were thawed in a water bath for 15 min at 37°C and mixed by pipetting before combining 216 µL urine with 9 µL of 1 M Tris-HCl to a final concentration of 40 mM. The samples, in two 100 µL volumes, were desalted by buffer exchange using a Zeba^TM^ 7 kDa MWCO Spin column desalting plate (ThermoFisher Scientific, Waltham, MA) according to the manufacturer’s guidelines. Desalting plates reached room temp before centrifuging at 1000 *x g* for 2 min to remove the storage buffer. A 250 µL volume of assay buffer was added to the resin beds and centrifuged at 1000 *x g* for 2 min. This was repeated three times. Protein concentration was determined using a Pierce^TM^ Micro BCA^TM^ protein assay kit (ThermoFisher Scientific, Waltham, MA) according to the manufacturer’s instructions.

#### SomaScan^®^ proteomic assay

The differential abundance of proteins in urine samples was measured using an aptamer-based SomaScan^®^ 7,000 protein microarray assay kit v4.1 (SomaLogic^®^, Boulder, CO, USA) as previously described [22]. Briefly, the microarray screened proteins using “Slow Off-Rate Modified Aptamers” (SOMAmers^®^) that were selected against protein epitopes using the Systematic Evolution of Ligands by Exponential Enrichment (SELEX) [22]. The scan utilized a resolution of 5 µm and detected Cy3 fluorescence expressed as relative fluorescence units (RFU). Off-scanner raw signal values were calibrated, standardized, and scaled at multiple dilution categories, namely S1 (61% of proteins), S2 (∼38.5%), and S3 (0.4%), where S1 targets low abundant proteins, and S3 targets proteins close to saturation. RFU readouts were analyzed using the Agilent Feature Extraction v10.7.3.1 (Agilent Technologies, Santa Clara, CA). Differential abundance was calculated using the SomaScan^®^ statistical analysis tool v4.1 and fitted as a linear model of the signal data and an empirical analysis by a Bayesian statistical test for group comparisons.

#### ELISA

Proteins selected for ELISA validation included REG1B (EH389RB, ThermoFisher Scientific, Waltham, MA), DEFA5 (HUFI01588, AssayGenie, Dublin, Ireland), FABP2 (EHFABP2, ThermoFisher Scientific, Waltham, MA), and REG3A (EH390RB, ThermoFisher Scientific, Waltham, MA). Urine samples were prepared, desalted, and quantified as described above. For each sample, triplicate measurements of 0.2, 2, 10, or 20 µg total protein were plated based on anticipated protein abundance to fit within the range of the standard curve for each ELISA.

#### Ingenuity Pathway Analysis

Network analysis and pathway enrichment were predicted using Ingenuity Pathway Analysis (IPA) (QIAGEN digital insights, Redwood City, CA, USA, accessed on June 20, 2023). IPA was used to generate a core analysis using all 99 differentially abundant proteins (**Table 2**) based on differential median fold change expression (Log_2_ ± 1.1) and Mann-Whitney U-test significance (*P* < 0.05) cut-offs.

**Table 2.**
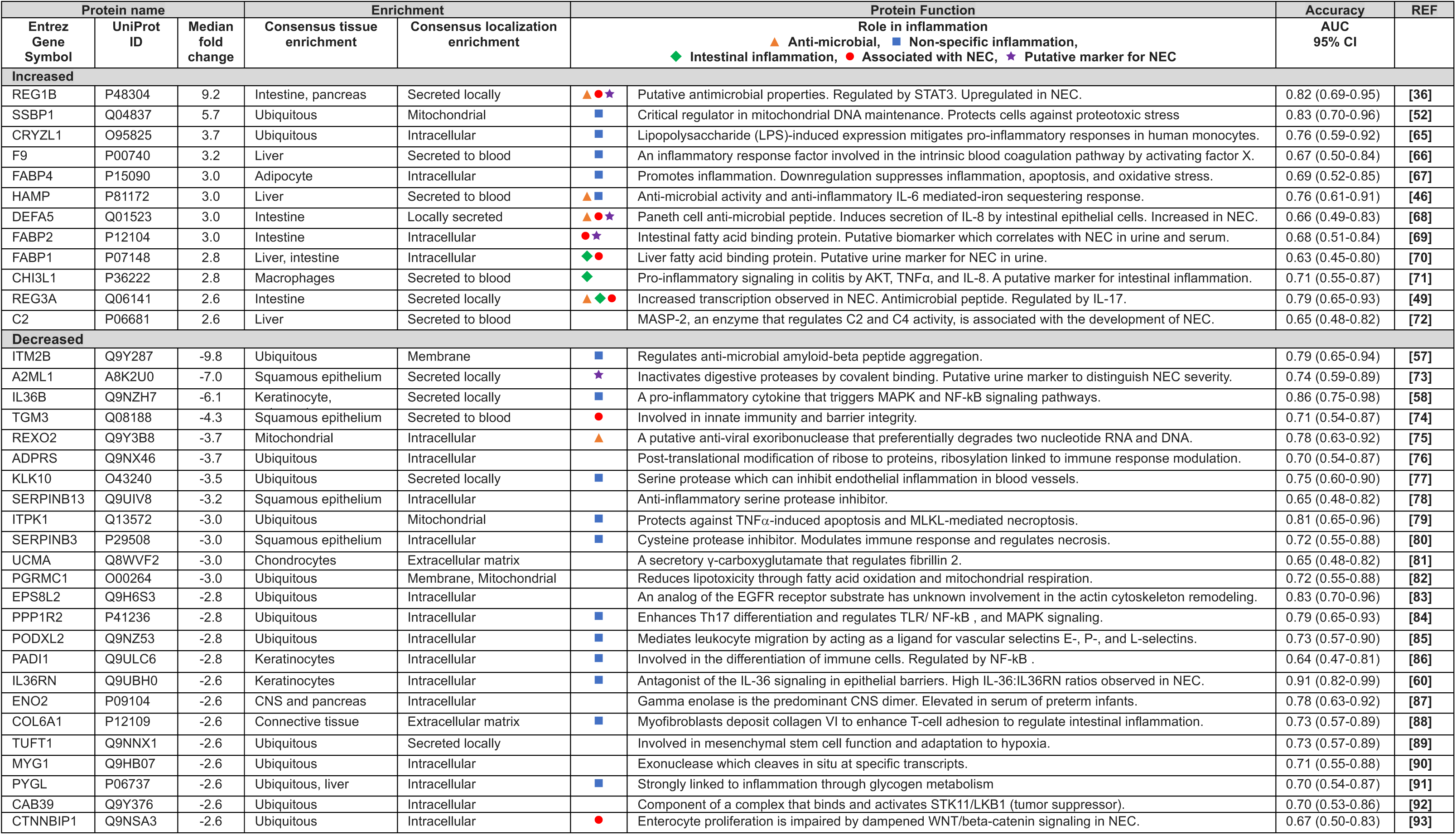
Classification of differentially abundant urine proteins identified by the SomaScan® (7K) assay.

#### Statistical analysis

Proteins above a Log_2_ median fold change RFU of 1.1 and a statistical significance of *P* < 0.05 for the comparison between samples from infants with NEC and controls were selected for detailed analysis. Samples were analyzed for the influence of multiple comparisons using a false discovery rate (FDR) of < 0.05. Volcano plots, hierarchical cluster analysis, and Spearman’s *r* correlation matrices were performed using Log_2_ RFU median fold change values. Log_10_ RFU values were used for comparative statistical analysis for violin plots and receiver operator characteristics (ROC). Statistical significance was determined by a Mann-Whitney U-test, non-parametric Wilcoxon test, or Kruskal Wallis as indicated. Combined ROC curves were generated using multiple logistic regression. Comparative statistical analysis was performed using GraphPad 9.3.1 (San Diego, USA). Hierarchical clustering was generated in R-studio version 4.2.0 using the “heatmap.2” package by Ward clustering.

## Results

### Cohort characteristics

The cohort included infants (*n* = 28) born between 24 and 36 weeks gestation and divided into age- and self-matched cohorts (**Table 1**). For the age-matched cohort, infants were enrolled at the time of NEC diagnosis (*n* = 8) and matched with infants who did not develop NEC (controls, *n* = 8). For the self-matched cohort, infants were initially enrolled as controls but then developed NEC (*n* = 12). Samples obtained before NEC diagnosis were used as the control for this group, with six days being the shortest interval between a control sample and a diagnosis of NEC.

### Qualitative and quantitative proteomic analysis of urine proteins

A total of 7596 proteins were screened in the urine samples using an aptamer-based proteomics microarray (SomaScan^®^). For the age-matched cohort, 150 urine proteins were significantly different between the infants with NEC and controls, based on Log_2_ median fold change RFU of ± 1.1 and Mann-Whitney U-test of *P* < 0.05 (**Figure 1A**). For the self-matched cohort, 360 urine proteins were differentially abundant in the control compared to the NEC samples (**Figure 1B**). There were 99 proteins shared between the age- and self-matched groups, and these will be the focus of the remainder of the analysis (**Figure 1C**). Of these 99 proteins, 35 were increased, and 64 were decreased between infants with NEC and their respective controls. A summary of the identified proteins in the combined cohort is provided in **Table S1**. Due to the small sample size of the cohort, only 10 proteins of the 7596 in the microarray survived multiple testing correction by false discovery rate (FDR < 0.05). Of the proteins that survived this correction, interleukin 36B (IL36B, FDR = 0.024) and interleukin 36 receptor antagonist (IL36RN, FDR = 0.024) were in the subset of proteins that were significantly different between the infants with NEC and controls.

**Figure 1.**
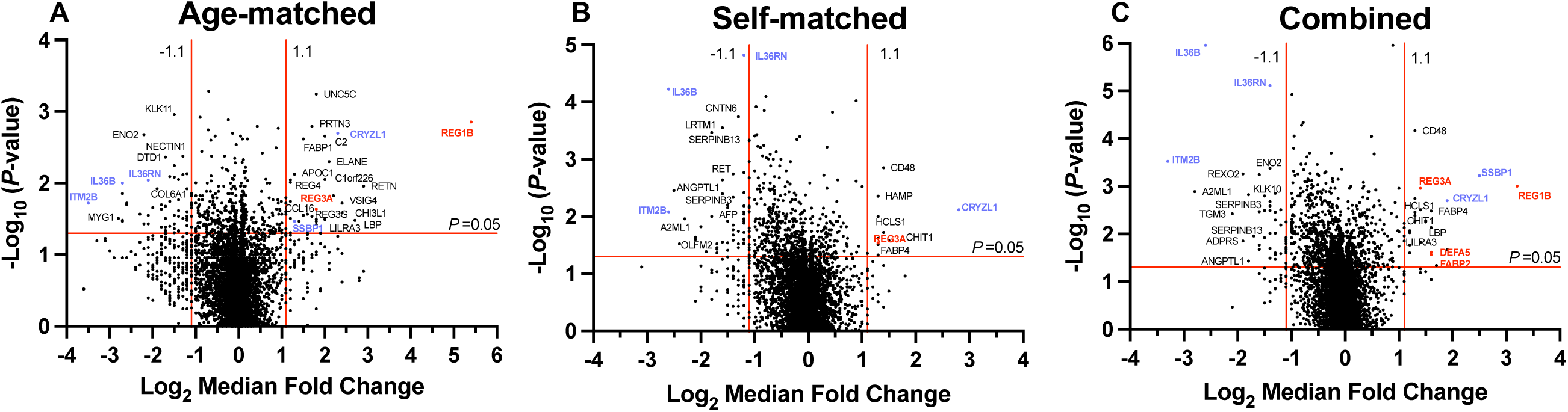
Volcano plots depicting the relative abundance of urine proteins in infants with and without NEC. Urine protein levels were compared between infants with and without NEC in the (**A**) Age-matched cohort (*n* = 8 pairs), (**B**) Self-matched cohort (*n* = 12 pairs), or (**C**) both groups combined (*n* = 20 pairs). Differentially abundant proteins are presented on a volcano plot with a median fold change of RFU values (Log_2_ ± 1.1) and *P*-value (−Log_10_ Mann Whitney U-test, *P* = 0.05) depicted as red lines. Proteins of interest are labeled red (Protein panel 1) or blue (Protein panel 2). REG1B was included in both panels.

We next identified two protein panels of interest for further analysis. Protein panel one consisted of four proteins chosen for their primary localization within the intestine and previously observed association with NEC and/or inflammation (**Table 2**). These include regenerating family member 1 beta (REG1B), regenerating islet-derived protein 3 alpha (REG3A), intestinal fatty acid binding protein 2 (FABP2), and defensin 5 alpha (DEFA5). Protein panel 2 consisted of proteins chosen based on the magnitude of the fold change between patients with NEC and controls, as well as the best area under the curve (AUC) values (**Table 2**). Panel 2 included REG1B (also in panel 1), single-stranded DNA binding protein 1 (SSBP1), crystallin zeta-like 1 (CRYZL1), integral membrane protein 2B (ITM2B), IL36B, and IL36RN.

We analyzed the association between the relative protein levels for the most differentially abundant proteins using an inter-protein Spearman’s *r* correlation matrix (**Figure 2A**). With a focus on our protein panels, we found that for panel 1, DEFA5 and REG3A had the strongest correlation (*r* = 0.83) (**Figure 2B**). In addition, REG3A (*r* = 0.57) and DEFA5 (*r* = 0.46) had strong positive correlations to REG1B (**Figure 2B**). For protein panel 2, there was a strong negative correlation with IL36RN with REG1B (*r* = -0.63) (**Figure 2C**). In addition, SSBP1 correlated positively with ITM2B (*r* = 0.62) and negatively with CRYZL1 (*r* = -0.52) and IL36B (*r* = -0.45) (**Figure 2C**).

**Figure 2.**
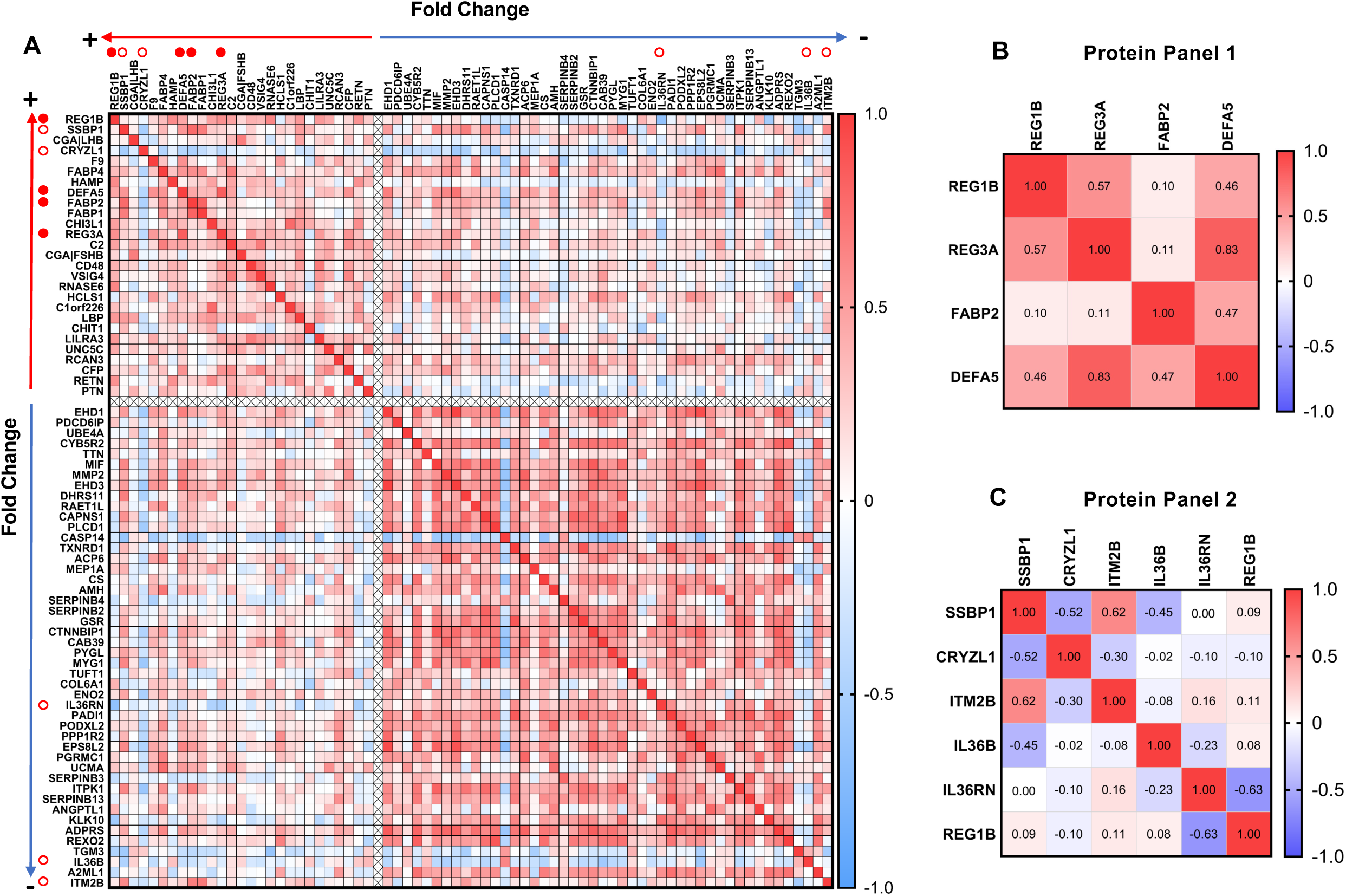
Correlation between relative protein levels in the urine of infants with and without NEC. (**A**) Spearman’s *r* correlation was used to determine the correlation of relative protein abundance for the entire cohort (*n* = 20 pairs). The directionality of the median fold change for NEC vs. non-NEC is depicted by red (increased) and blue (decreased) arrows. Protein panel 1 (⚫). Protein panel 2 (○). REG1B was included in both panels. Correlation matrices were generated for protein panels 1 (**B**) and 2 (**C**).

We next used hierarchical cluster analysis to visualize patterns in protein abundance across patient pairs and disease severity (**Figure 3**). We found that the pattern of relative fold change of the proteins was consistent across samples. In addition, age-matched and self-matched samples did not cluster together, but samples did cluster based on disease severity, with noted clustering of samples from patients with Bell’s stage III NEC (**Figure 3**).

**Figure 3.**
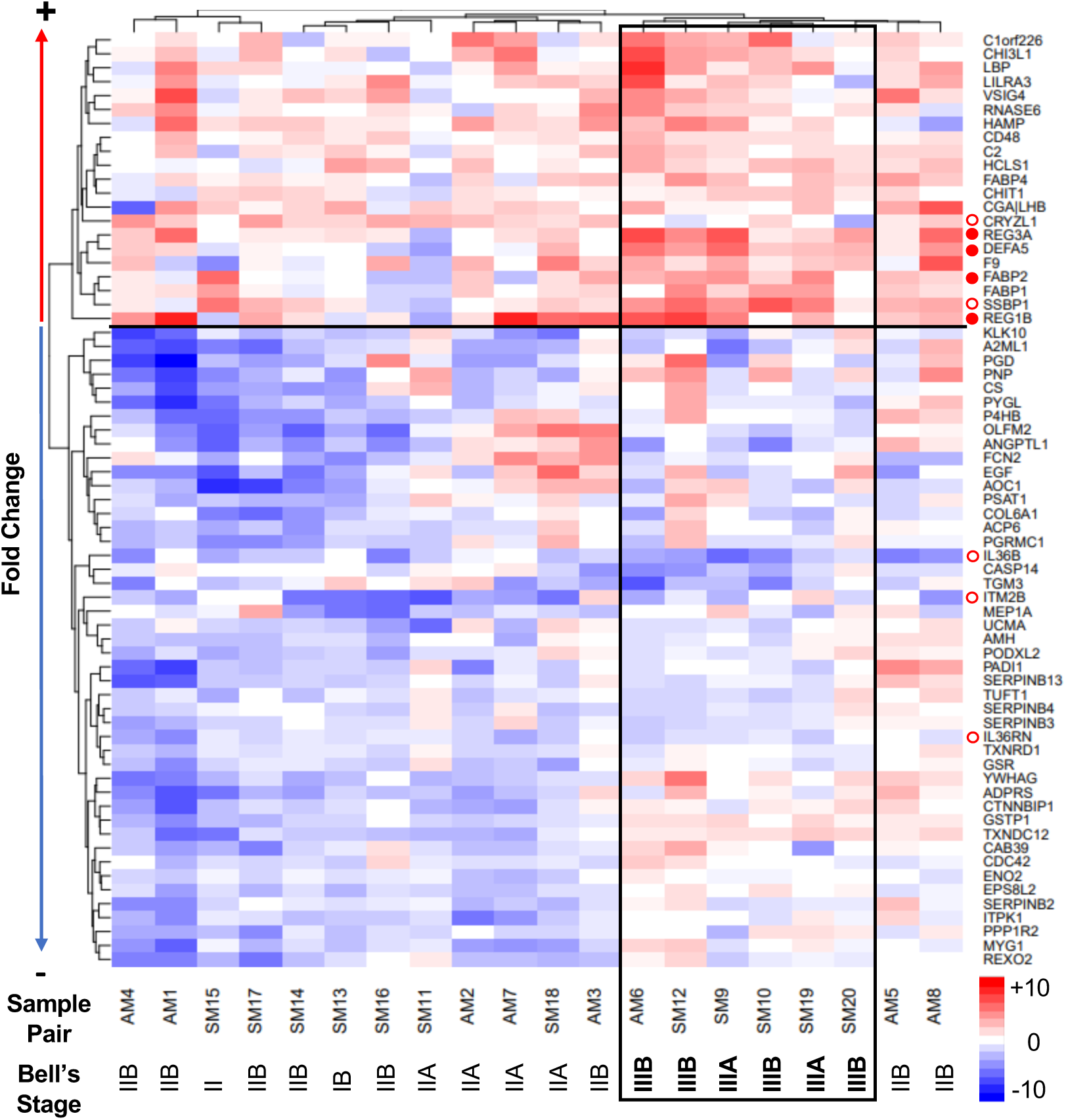
Relative protein levels in matched urine samples for pairs of infants with and without NEC cluster based on Bell’s Stage, a marker of NEC severity. Hierarchical cluster analysis demonstrating median fold change in protein abundance for sample pairs. The directionality of the median fold change for NEC vs. control is depicted by red (increased) and blue (decreased) arrows. Protein panel 1 (⚫). Protein panel 2 (○). The black box delineates patients with Bell’s Stage III NEC.

### Analysis of protein abundance and comparison based on disease severity

Violin plots were used to visualize the protein abundance in urine samples for the proteins in panel 1, with significant differences noted between controls and patients with NEC (**Figure 4A**). We further analyzed the differences in urine protein abundance based on disease severity as defined by modified Bell’s stage (**Figure 4B**). REG1B was significantly increased in infants with NEC Bell’s stage II compared to controls (*P* < 0.01) as well as in Bell’s stage III vs. controls (*P* < 0.01). REG3A and DEFA5 were significantly increased in the urine of infants with NEC Bell’s stage III vs. controls (REG3A: *P* < 0.0001, DEFA5: *P* < 0.0005) and in Bell’s stage II compared to Bell’s stage III (REG3A: *P* < 0.01, DEFA5: *P* < 0.01). FABP2 was only significantly increased Bell’s stage III vs. controls (*P* < 0.05).

**Figure 4.**
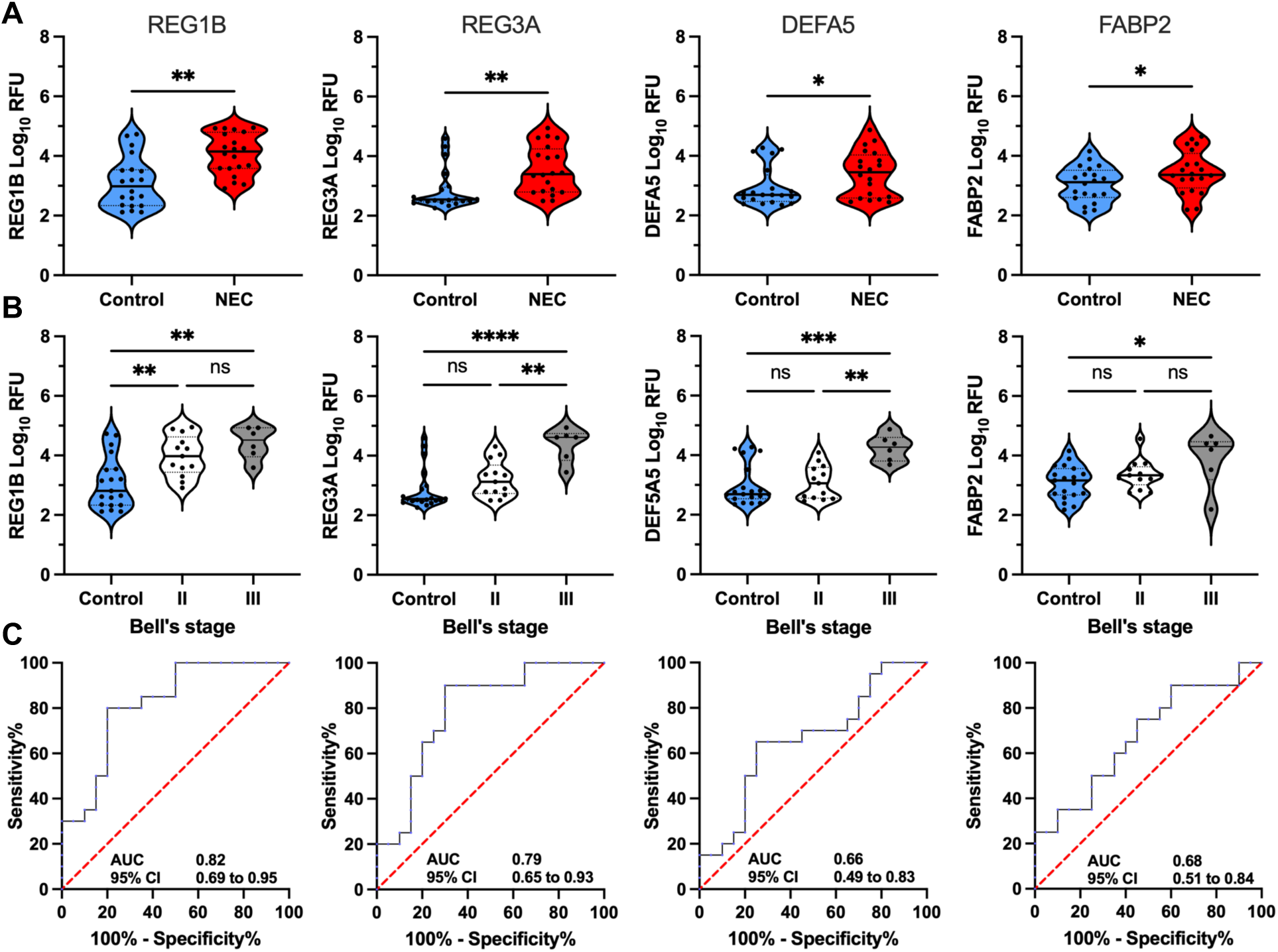
Urine proteins in protein panel 1 differ between infants with NEC and controls, as well as based on Bell’s stage. The proteins in panel 1 include REG1B, REG3A, DEFA5, and FABP2. (**A**) Comparison of protein abundance for infants with NEC and controls. Line indicates median. Dots indicate individual infants. * *P* < 0.05, ** *P* < 0.01 by non-parametric Wilcoxon test. (**B**) Samples were stratified based on disease severity using modified Bell’s stage. Line indicates median. Dots indicate individual infants. ns, *P* > 0.05, * *P* < 0.05, ** *P* < 0.01, *\*\*\* P <* 0.0005, **** *P* < 0.0001 by Kruskal Wallis. (**C**) ROC curves with AUC values and 95% CI.

ELISAs were used to validate the relative protein abundance detected with the protein microarray. We selected the proteins in panel 1, REG1B, REG3A, FABP2, and DEFA5 (**Supplementary Figure S1**) for validation. REG1B (**Figure S1A**) and DEFA5 (**Figure S1C**) were both significantly increased in NEC relative to controls; however, REG3A (**Figure S1B**) and FABP2 (**Figure S1D**) were not significantly different in NEC compared to controls (**Supplementary Table S2**). This likely occurred because levels of the urine proteins were below the limit of detection for the ELISAs for some patients, which resulted in an inability to detect differences between patients with NEC and controls. These assays were limited by the volume of urine obtainable from preterm neonates and the low concentration of the proteins of interest. Thus, the microarray data was used for the remainder of the analysis due to increased sensitivity at lower protein concentrations.

Analysis of the ability of these proteins to serve as potential biomarkers was performed by calculating the AUC (**Table 2** and **Figure 4C**). Values above 0.7 are considered acceptable, and above 0.8 are considered excellent for a diagnostic test. REG1B (0.82, 95% CI 0.69-0.95) and REG3A (0.79, 95% CI 0.69-0.95) were the most effective at differentiating between patients with NEC and controls (**Figure 4C**).

We performed a similar analysis for the proteins in panel 2. The RFU data for the urine samples from patients with NEC and controls is shown in **Figure 5A**. We also analyzed the ability of these proteins to differentiate between patients based on NEC severity (**Figure 5B**). We found that CRYZL1, ITM2B, IL36B, and IL36RN were significantly different between infants with Bell’s stage II NEC relative to controls (*P* < 0.05). SSBP1 (*P* < 0.005) and IL36B (*P* < 0.05) were significantly different between infants with NEC Bell’s Stage II and III.

**Figure 5.**
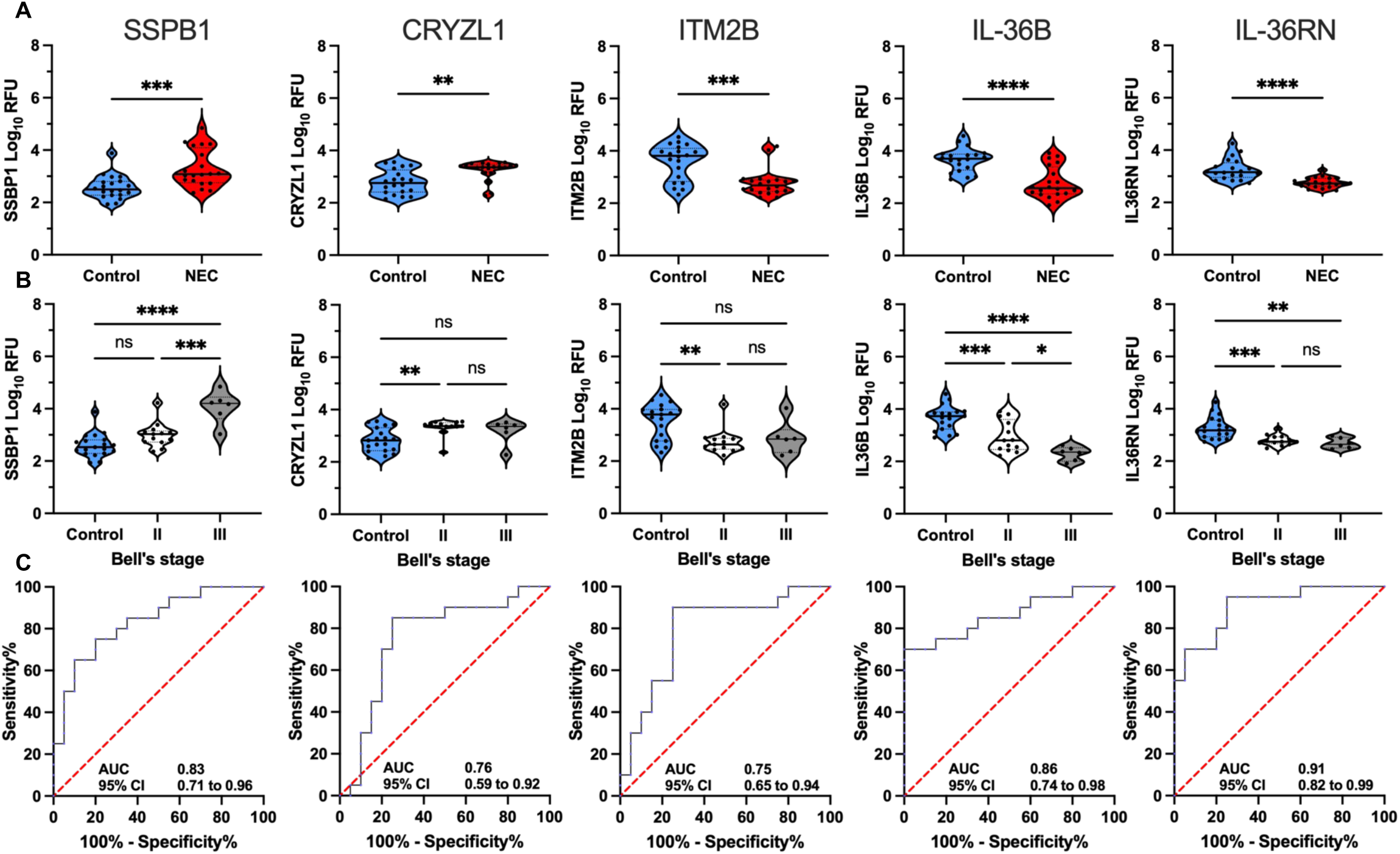
Urine proteins in protein panel 2 differ between infants with NEC and controls, as well as based on Bell’s stage. (**A**) Comparison of protein abundance for infants with NEC and controls. Line indicates median. Dots indicate individual infants. * *P* < 0.05, ** *P* < 0.01, *** *P* < 0.005, **** *P* < 0.0001 by non-parametric Wilcoxon test. (**B**) Samples were stratified based on disease severity using modified Bell’s stage. Line indicates median. Dots indicate individual infants. * *P* < 0.05, ** *P* < 0.01, *** *P* < 0.005 by Kruskal Wallis. (C) ROC curves with AUC values and 95% CI.

ROC curves were generated for all proteins in panel 2. All of the proteins in panel 2 were effective at differentiating between infants with NEC and controls (**Figure 5C**). IL36B (0.86, 95% CI 0.74-0.98) and IL36RN (0.91, 95% CI 0.82-0.99) had the best AUC values and have the potential to serve as excellent diagnostic tests.

### Diagnostic utility of protein panels

We generated combined ROC curves for groups of proteins to determine if this would improve their diagnostic utility as urine biomarkers (**Figure 6**). We found that protein panel 1 (**Figure 6A**, AUC 0.9, 95% CI 0.8-1), protein panel 2 (**Figure 6B**, AUC 0.98, 95% CI 0.95-1), and the combination of IL-36B and IL36RN (**Figure 6C**, AUC 0.94, 95% CI 0.86-1) had an outstanding ability to differentiate between infants with NEC and controls.

**Figure 6.**
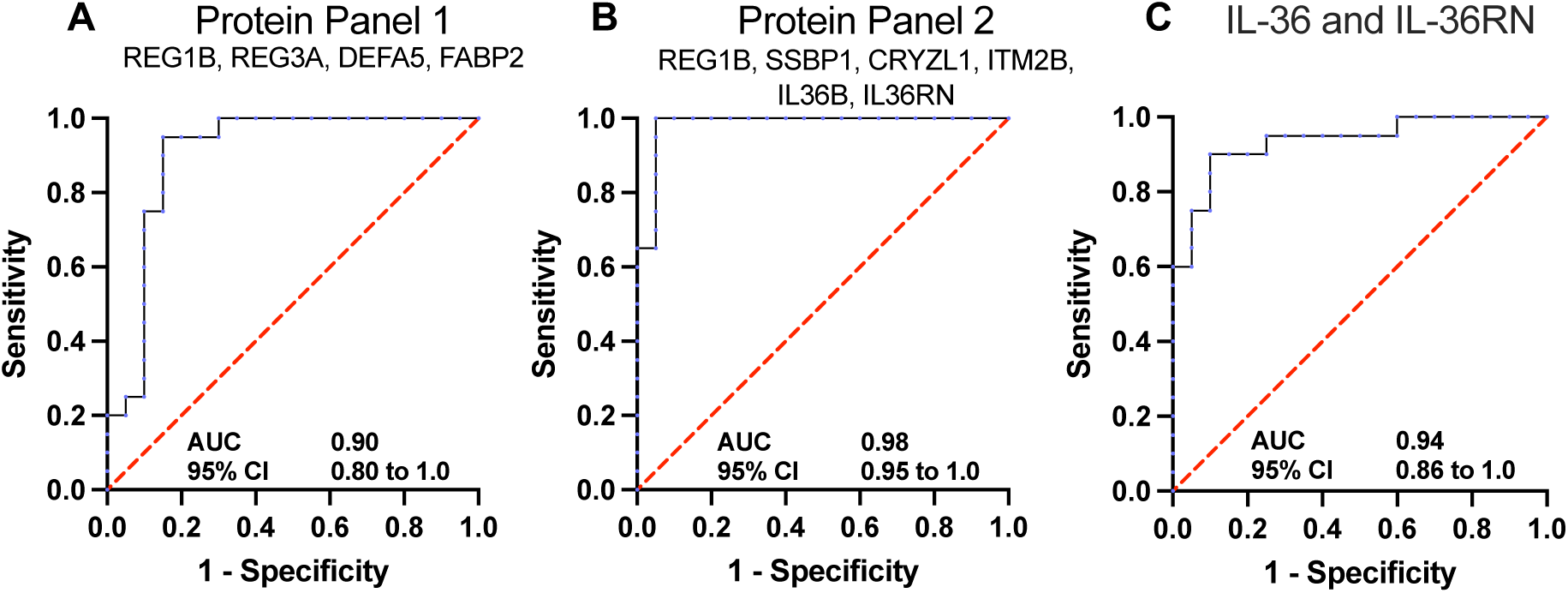
Combinations of select urine proteins very effectively discriminate between infants with and without NEC. Combined ROC curves were generated for (**A**) Protein panel 1 (REG1B, REG3A, DEFA5, and FABP2), (**B**) Protein panel 2 (REG1B, SSBP1, CRYZL1, ITM2B, IL36B, and IL36RN) and (**C**) IL-36 and IL36RN. AUC values with the 95% CI are shown.

### Upstream regulators and statistical enrichment

Ingenuity pathway analysis (IPA) was used to predict activated or inhibited upstream regulators and signaling pathways. IPA predicts differentially activated regulators measured by downstream enrichment scores (overlap *P*-value) and activation states (activation *z*-score) [23]. Upstream regulator analysis (URA) predicts direct relationships between upstream regulators and downstream molecules using the protein dataset. Causal analysis (CNA) tools further predict upstream master regulators that may have multiple indirect relationships, nodes, and links. Upstream regulators of interest (linked to inflammation) by URA and CNA were predicted and summarized in **Table 3**.

**Table 3.**
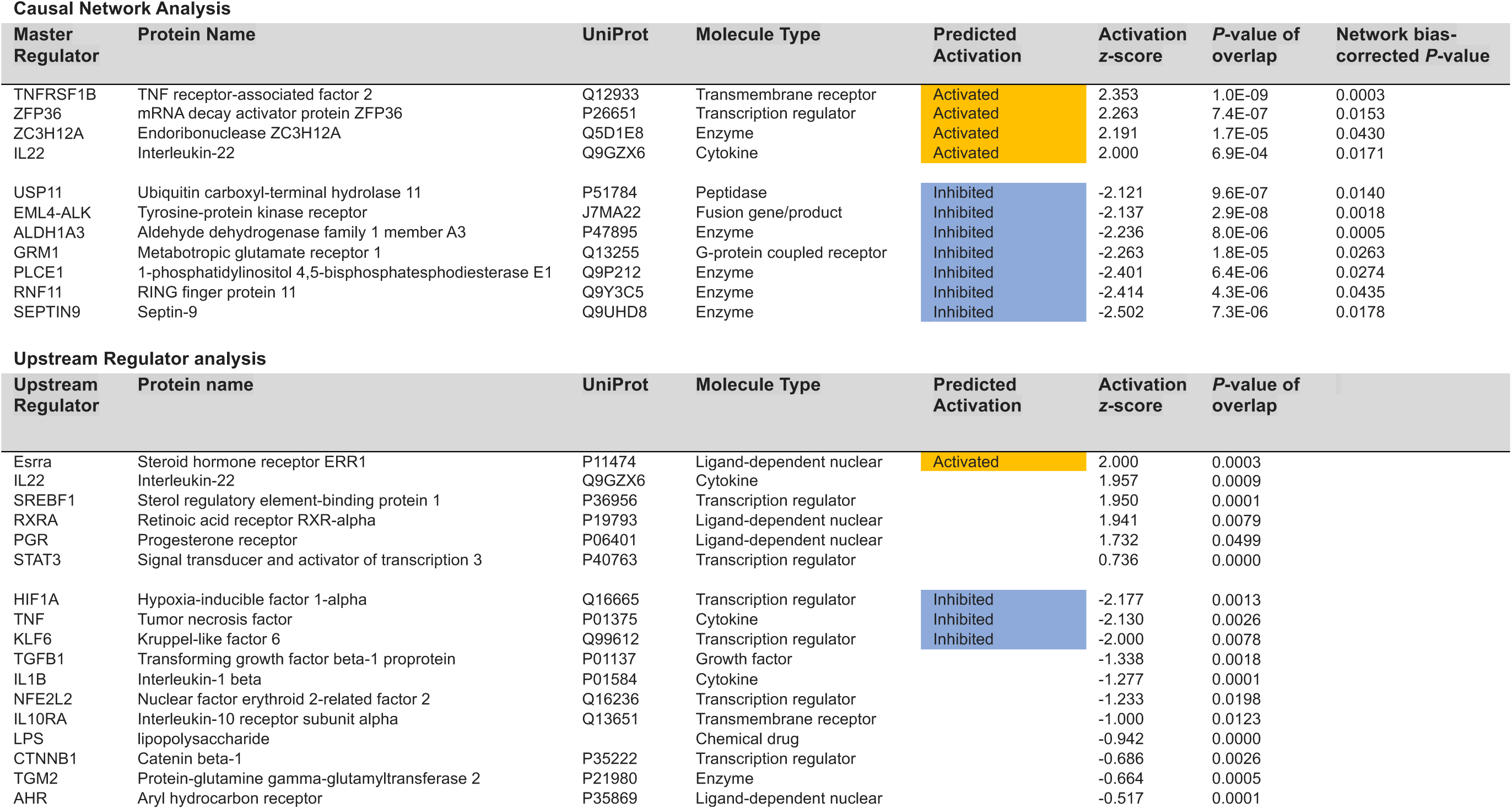
Ingenuity pathway upstream regulator analysis

URA predicted several direct regulators based on the downstream status of the differentially abundant proteins identified in the urine samples. This includes regulators of lipid metabolism, sterol regulatory element binding protein 1 (SREBF1, *z* = 1.95, *P* = 0.00015), and retinoic acid receptor alpha (RXRA, *z* = 1.94, *P* = 0.00794) (**Table 3**). In addition, the polyfunctional cytokine IL-22 (*z* = 1.96, *P* = 0.00091) was predicted as activated based on the differential abundance of REG1B (+), REG3A (+), LBP (+), and TGM3 (-) (**Supplementary Figure S2, Table 3**).

CNA includes indirect relationships and linked nodes to identify upstream-master regulators (**Table 3**). Predicted master regulators included: TNF receptor-associated factor 1B (TRAF2) (*z* =2.353, network bias corrected P = 0.0003), which plays a role in NFκB and JNK signaling pathways [24], mRNA decay activator protein ZFP36 (*z* = 2.263, network bias corrected *P* = 0.0153), which is involved in cytokine degradation and regulates the TNF-alpha feedback loop through mRNA destabilization [25,26], Endoribonuclease ZC3H12A (*z* = 2.191, network bias corrected P = 0.043), and IL-22 (*z* = 2.0, network bias corrected *P* = 0.0171). STAT3, which is activated by IL-22 and other cytokines, was upstream of several differentially abundant proteins in this study (**Supplementary Figure S2, Table 3**).

## Discussion

In this study, we used an aptamer-based protein microarray to successfully identify a subset of urine proteins that differentiated between infants with and without NEC. The validity of our data was supported by the fact that patterns of relative protein abundance in the urine were consistent across both age- and self-matched cohorts. In addition, the protein levels in the urine of these patient cohorts clustered based on the severity of NEC. Our identified biomarker panels had excellent diagnostic potential based on AUC values, supporting further studies investigating these proteins of interest as potential biomarkers.

Our study focused on urine as the biological source for biomarker testing, given that it is an easily obtainable specimen even for tiny preterm neonates. Unfortunately, the detection of biomarkers in urine adds additional analytical challenges due to sample-to-sample biological variability. This includes significant variability in hydration states, glomerular filtration rates (GFR), and differential abundance in carrier proteins and metabolites [27]. The accurate quantitation of differentially abundant proteins measured in urine is likely improved using a reference marker. Typical urine reference markers require a constitutively expressed gene or metabolite unaffected by the disease. Studies have tried to identify a reliable normalization reference using urinary proteins or peptides to adjust for urinary dilution [7]. These have included serum albumin, collagen, and urine creatinine; however, protein and metabolite variability based on the developmental age of neonates complicates normalization [28]. For example, urine albumin and creatinine concentrations in neonates have been shown to be inconsistent in preterm infants [29]. The consistency of our protein abundance patterns with two different patient matching strategies and across a range of corrected gestational ages indicates that these technical issues were likely minimized by using equivalent protein loading, as was performed in our studies.

Based on published data, the likely intestinal origin of many of the proteins we detected provides another degree of complexity, as it requires proteins or peptides to enter the bloodstream from the intestine and be filtered by the glomeruli in the kidney to be identified in the urine. In addition, biomarkers need to distinguish NEC from non-specific inflammation in the kidneys and urinary tract to have clinical significance. In this study, known biomarkers for urinary tract infections [30] and acute kidney injury [31], specifically NGAL (Lipocalin-2) and CST3 (Cystatin C), were not significantly abundant. In addition, the consistency of protein abundance patterns across matching strategies and the clustering of patients based on disease severity supports a NEC-specific protein signature independent of renal complications.

In this study, we developed two protein panels and identified a pair of proteins with excellent AUC values for diagnostic studies. Using a protein panel increases the power of a diagnostic test to accurately discriminate between healthy and diseased states. Although a single disease biomarker would be optimal, this is likely unfeasible given the complexity of the pathogenesis of NEC. Below, we discuss the specific proteins we incorporated into our panels.

### REG1B and REG3A

Regenerating islet-derived proteins (REGs) have multifunctional roles in cellular processes, including promoting differentiation, increasing cellular proliferation, preventing apoptosis, and enhancing host defense [32]. REG proteins (18-19 kDa) have been discovered in multiple tissues and diseases, and they are typically expressed at sites of inflammation [33]. REG1B and REG3A recognize bacterial peptidoglycan and reduce bacterial membrane integrity [34]. They are increased in inflammatory bowel disease (IBD), specifically ulcerative colitis (UC) [35]. REG1B expression is also increased in the intestine during NEC [36]. In addition, increased REG1B expression drives intestinal crypt regeneration [37]. REG3A has anti-apoptotic effects and increases enterocyte viability by reducing caspase cleavage [38]. REG1B [39] and REG3A [40] have been identified as potential stool biomarkers for enteropathy and environmental enteric dysfunction (EED). Finally, REG1B and REG3A are induced by the IL-22 signaling pathway [41], which is protective in the intestine of mice during experimental NEC [42].

### FABP2

Fatty acid binding protein 2 (FABP2) is a 15 kDa enterocyte-specific carrier protein involved in intracellular fatty acid translocation across the gut barrier [43]. FABPs are subdivided into intestinal (FABP2), hepatic (FABP1), and adipose (FABP4) variants based on localization.

FABP2, FABP1, and FABP4 were all differentially abundant in our proteomic assay (**Supplementary Table 1**). FABP2 has previously been identified as differentially abundant during intestinal barrier dysfunction [44] and, specifically, NEC in several biomarker studies of stool and urine [45]. A clinical metadata study showed a high FABP2 level was an indicator of NEC, but many studies were limited in sample size and had difficulty consistently normalizing urine samples by creatinine [12].

### DEFA5

Human α-defensin (DEFA5) (10 kDa) is a powerful antimicrobial host defense protein [46]. It is primarily expressed by intestinal Paneth cells and stored intracellularly as a zymogen. After secretion, DEFA5 is activated by trypsin and regulated by Paneth cell-derived serine protease inhibitors, which play a role in selectively maintaining gut microbiome diversity. One study has linked increased expression with urinary tract infections [47]. Increased DEFA5 has previously been detected in the stool of infants with NEC via proteomics [48] and in intestinal samples resected from infants with NEC via RNA sequencing [49]. In addition, altered stool *DEFA5* methylation patterns have been observed during NEC [50].

### SSBP1

Single-stranded DNA binding protein 1 (SSBP1) is a ubiquitously expressed protein involved in mitochondrial biogenesis. Mitochondrial damage, through the presence of mitochondrial DNA and proteins in urine, has been used as a marker for tissue damage during inflammation [51]. In addition, SSBP1, through heat shock factor 1 (HSF1) signaling, has been linked to increased expression of heat shock protein 70 (Hsp70) [52]. Hsp70 protects cells from proteolytic stress and regulates pro-inflammatory Toll-like receptor (TLR)-induced cytokines [53]. In enterocytes, activation of TLR4 leads to upregulation of Hsp70, and Hsp70 signaling has been identified as a potential counter-regulatory mechanism that opposes TLR4 signaling during NEC [54].

### CRYZL1

Crystallin zeta-like 1 (CRYZL1) is ubiquitously expressed and shares sequence homology with zeta crystallin or quinone oxidoreductase 1 (NQO1). While the role of CRYZL1 is unclear, NQO1 protects against cellular damage by acting as a redox switch interacting with multiple proteins, mRNA, and NADH [55]. Upregulation of *Nqo1* expression has been associated with protection from NEC-like injury in rats [56].

### ITM2B

Integral membrane protein 2B (ITM2B) is found in all tissue types. Its exact function needs to be delineated, although it is thought to play a role in apoptosis, mitochondrial homeostasis, and processing of amyloid precursor proteins [57].

### IL36B and IL36RN

Interleukin 36 (IL-36) proteins are a family of inflammatory cytokines and consist of agonists IL36 (α, β, γ) and a receptor antagonist, IL36RN (IL36Ra). IL-36 dysregulation is associated with inflammatory diseases, including psoriasis, arthritis, and IBD [58]. In addition, IL-36 regulates both immune and non-immune cells in a feedback loop where IL-22, IL-17A, and TNFα induce all three IL-36 family cytokines, and IFNγ is selective for inducing IL36B [59].

Interestingly, in mice with NEC, increased IL36B was detected only in the ileum, and an increased IL36B: IL36RN ratio was associated with NEC [60]. In our study, the relative decrease in IL36RN is consistent with increased inflammation, whereas the reduction in IL36B is not expected and needs further exploration in mechanistic studies.

### Upstream regulator network analysis

IPA identified several potential upstream and mechanistic regulators. Notably, IL-22 was predicted to be an upstream regulator based on the downstream differential abundance of REG1B, REG3A, LPS-binding protein (LBP), and transglutaminase 3 (TGM3). IL-22, through STAT3-dependent signaling [61], is involved in intestinal barrier protection and induces proliferation in intestinal stem cells and goblet cell proliferation in the crypts [62]. Of note, STAT3 is activated by cytokines other than IL-22, such as IL-6 and IL-10 family cytokines [63]. The redundancy in these pathways makes it difficult to pinpoint the upstream mediators without mechanistic data; however, the increased abundance of these intestinal proteins in the urine of infants with NEC indicates a potential increase in signaling via STAT3 activating cytokines and/or is reflective of a weakened gut barrier in patients with NEC. Additionally, hepcidin (HAMP), which had a 3-fold increased abundance in the samples from infants with NEC, is an antimicrobial iron sequestering protein and is also positively regulated by IL-22 [64]. Moreover, IL-22 is a complex and polyfunctional cytokine. We have previously shown that administration of recombinant IL-22 is protective against intestinal injury during experimental NEC in mice [42]; however, further studies are needed to determine the balance between protective and inflammatory roles for this cytokine in NEC.

## Conclusion

Due to the rapid and unpredictable onset of NEC, a biomarker needs to be readily available, non-invasive, rapidly tested, and able to identify NEC prior to the advanced Bell’s stages. In this study, we identify proteins of interest that have the potential to serve as biomarkers that are so urgently needed to improve the care of infants with NEC. While our results need to be validated in a large cohort of infants with diverse physiologic and inflammatory conditions, this study will serve as an important foundation for future studies on a larger, diverse cohort.

## Declarations

### Ethics approval and consent to participate

After consent was obtained, samples were prospectively collected from infants admitted to the St. Louis Children’s Hospital Neonatal Intensive Care Unit (NICU) in St. Louis, Missouri, USA. Enrollment was conducted according to protocols approved by the Washington University in St. Louis School of Medicine Institutional Review Board (IRB protocol numbers 201706182 and 201802101).

## Availability of data and materials

All data generated or analyzed during this study are included in this published article and its supplementary information files.

## Authors contributions

Manuscript preparation by SM, LF, CM, and MG. Critical review of the manuscript by all authors. Data analysis by SM and LF. Research coordination and sample processing by QG, OD, GB, and MG. Study design by SM, LF, and MG.

## Data Availability

All data produced in the present work are contained in the manuscript and are available upon reasonable request to the corresponding author.

## Acknowledgements

We want to thank the neonates and families who participated in our study. We would also like to thank the staff at St. Louis Children’s Hospital Neonatal Intensive Care Unit in St. Louis, Missouri, USA, for all their hard work in helping to collect samples, which made this study possible.

---

**Supplementary Figure S1. ELISA validation of urine protein levels for controls and infants with NEC.** (**A**) REG1B, (**B**) REG3A, (**C**) DEFA5, and (**D**) FABP2 were measured in the urine of all infants in the study via ELISA. Samples with protein levels below the limit of detection were censored. Bar indicates median. n = number of patients with detectable urine protein. ns*, P* > 0.05, * *P* < 0.05, ** *P* < 0.01 via Mann Whitney U-test.

**Supplementary Figure S2. IL-22 and STAT3 are upstream regulators of proteins that are differentially abundant in the urine of infants with and without NEC.** (**A**) Upstream regulator analysis (URA) predicted IL-22 as a direct regulator based on the downstream state of REG1B (+), REG3A (+), LBP (+), and TGM3 (-). (**B**) Causal network analysis (CNA) mapped indirect relationships, which included IL-22 and STAT3. Proteins marked in red and green were increased and decreased, respectively. Orange and blue proteins and interactions indicate predicted activation and inhibition, respectively. Yellow and grey lines indicate interactions that are inconsistent or not predicted with the state of the downstream molecule. Dashed and solid lines indicate indirect and direct relationships, respectively.

## List of abbreviations

(NEC): Necrotizing enterocolitis
(ROC): receiver operator characteristics
(AUC): area under the curve
(RFU): relative fluorescence units
(FDR): false discovery rate
(IPA): Ingenuity pathway analysis
(URA): Upstream regulator analysis
(CNA): Causal analysis

## References

[1] Battersby C, Santhalingam T, Costeloe K, et al. Incidence of neonatal necrotising enterocolitis in high-income countries: a systematic review. Arch Dis Child Fetal Neonatal Ed. 2018 Mar;103(2):F182–f189.

[2] Goldstein GP, Sylvester KG. Biomarker Discovery and Utility in Necrotizing Enterocolitis. Clinics in Perinatology. 2019 2019/03/01/;46(1):1-17.

[3] Nagpal R, Tsuji H, Takahashi T, et al. Gut dysbiosis following C-section instigates higher colonisation of toxigenic Clostridium perfringens in infants. Benef Microbes. 2017 May 30;8(3):353–365.

[4] Adams JM, Valentine CJ, Karns RA, et al. DHA Supplementation Attenuates Inflammation-Associated Gene Expression in the Mammary Gland of Lactating Mothers Who Deliver Preterm. J Nutr. 2022 Jun 9;152(6):1404–1414.

[5] Sampah MES, Hackam DJ. Dysregulated Mucosal Immunity and Associated Pathogeneses in Preterm Neonates. Front Immunol. 2020;11:899.

[6] Singh DK, Miller CM, Orgel KA, et al. Necrotizing enterocolitis: Bench to bedside approaches and advancing our understanding of disease pathogenesis [Review]. Frontiers in Pediatrics. 2023 2023-January-11;10.

[7] Wang K, Tao G, Sun Z, et al. Recent Potential Noninvasive Biomarkers in Necrotizing Enterocolitis. Gastroenterol Res Pract. 2019;2019:8413698.

[8] Wang K, Tao G, Sun Z, et al. Recent Potential Noninvasive Biomarkers in Necrotizing Enterocolitis. Gastroenterology Research and Practice. 2019 2019/04/22;2019:8413698.

[9] Agakidou E, Agakidis C, Gika H, et al. Emerging Biomarkers for Prediction and Early Diagnosis of Necrotizing Enterocolitis in the Era of Metabolomics and Proteomics. Front Pediatr. 2020;8:602255.

[10] Donda K, Torres BA, Maheshwari A. Non-coding RNAs in Neonatal Necrotizing Enterocolitis. Newborn (Clarksville). 2022 Jan-Mar;1(1):120–130.

[11] Galley JD, Mar P, Wang Y, et al. Urine-derived extracellular vesicle miRNAs as possible biomarkers for and mediators of necrotizing enterocolitis: A proof of concept study. Journal of Pediatric Surgery. 2021 2021/11/01/;56(11):1966–1975.

[12] Liu Y, Jiang L-F, Zhang R-P, et al. Clinical significance of FABP2 expression in newborns with necrotizing enterocolitis. World Journal of Pediatrics. 2016 2016/05/01;12(2):159–165.

[13] Chaaban H, Markel TA, Canvasser J, et al. Biobanking for necrotizing enterocolitis: Needs and standards. J Pediatr Surg. 2020 Jul;55(7):1276–1279.

[14] Ralls MW, Gadepalli SK, Sylvester KG, et al. Development of the necrotizing enterocolitis society registry and biorepository. Semin Pediatr Surg. 2018 Feb;27(1):25–28.

[15] Soomro S, Venkateswaran S, Vanarsa K, et al. Predicting disease course in ulcerative colitis using stool proteins identified through an aptamer-based screen. Nat Commun. 2021 Jun 28;12(1):3989.

[16] Vanarsa K, Castillo J, Wang L, et al. Comprehensive proteomics and platform validation of urinary biomarkers for bladder cancer diagnosis and staging. BMC Medicine. 2023 2023/04/05;21(1):133.

[17] Dong L, Watson J, Cao S, et al. Aptamer based proteomic pilot study reveals a urine signature indicative of pediatric urinary tract infections. PLoS One. 2020;15(7):e0235328.

[18] Billing AM, Ben Hamidane H, Bhagwat AM, et al. Complementarity of SOMAscan to LC-MS/MS and RNA-seq for quantitative profiling of human embryonic and mesenchymal stem cells. J Proteomics. 2017 Jan 6;150:86–97.

[19] Neu J, Modi N, Caplan M. Necrotizing enterocolitis comes in different forms: Historical perspectives and defining the disease. Semin Fetal Neonatal Med. 2018 Dec;23(6):370–373.

[20] Bell MJ, Ternberg JL, Feigin RD, et al. Neonatal necrotizing enterocolitis. Therapeutic decisions based upon clinical staging. Ann Surg. 1978 Jan;187(1):1–7.

[21] Walsh MC, Kliegman RM. Necrotizing enterocolitis: treatment based on staging criteria. Pediatr Clin North Am. 1986 Feb;33(1):179–201.

[22] Gold L, Ayers D, Bertino J, et al. Aptamer-based multiplexed proteomic technology for biomarker discovery. PLoS One. 2010 Dec 7;5(12):e15004.

[23] Krämer A, Green J, Pollard J, Jr., et al. Causal analysis approaches in Ingenuity Pathway Analysis. Bioinformatics. 2014 Feb 15;30(4):523–30.

[24] Lee SY, Reichlin A, Santana A, et al. TRAF2 is essential for JNK but not NF-kappaB activation and regulates lymphocyte proliferation and survival. Immunity. 1997 Nov;7(5):703–13.

[25] Kuiken HJ, Egan DA, Laman H, et al. Identification of F-box only protein 7 as a negative regulator of NF-kappaB signalling. J Cell Mol Med. 2012 Sep;16(9):2140–9.

[26] Carballo E, Lai WS, Blackshear PJ. Feedback inhibition of macrophage tumor necrosis factor-alpha production by tristetraprolin. Science. 1998 Aug 14;281(5379):1001–5.

[27] Jantos-Siwy J, Schiffer E, Brand K, et al. Quantitative urinary proteome analysis for biomarker evaluation in chronic kidney disease. J Proteome Res. 2009 Jan;8(1):268–81.

[28] Sylvester KG, Moss RL. Urine biomarkers for necrotizing enterocolitis. Pediatr Surg Int. 2015 May;31(5):421–9.

[29] Sanderson K, O’Shea TM, Kistler CE. An Individualized Approach to Kidney Disease Screening in Children With a History of Preterm Birth. Clin Pediatr (Phila). 2023 Jun;62(5):385–388.

[30] Horváth J, Wullt B, Naber KG, et al. Biomarkers in urinary tract infections - which ones are suitable for diagnostics and follow-up? GMS Infect Dis. 2020;8:Doc24.

[31] Vaidya R, Yi J, O’Shea T, et al. Long-Term Outcome of Necrotizing Enterocolitis and Spontaneous Intestinal Perforation. Pediatrics. 2022 10/06;150.

[32] Cash HL, Whitham CV, Behrendt CL, et al. Symbiotic bacteria direct expression of an intestinal bactericidal lectin. Science. 2006 Aug 25;313(5790):1126–30.

[33] Sun C, Wang X, Hui Y, et al. The Potential Role of REG Family Proteins in Inflammatory and Inflammation-Associated Diseases of the Gastrointestinal Tract. Int J Mol Sci. 2021 Jul 3;22(13).

[34] Mukherjee S, Hooper Lora V. Antimicrobial Defense of the Intestine. Immunity. 2015 2015/01/20/;42(1):28–39.

[35] van Beelen Granlund A, Østvik AE, Brenna Ø, et al. REG gene expression in inflamed and healthy colon mucosa explored by in situ hybridisation. Cell Tissue Res. 2013 Jun;352(3):639–46.

[36] Egozi A, Olaloye O, Werner L, et al. Single-cell atlas of the human neonatal small intestine affected by necrotizing enterocolitis. PLOS Biology. 2023;21(5):e3002124.

[37] Peterson KM, Guo X, Elkahloun AG, et al. The expression of REG 1A and REG 1B is increased during acute amebic colitis. Parasitol Int. 2011 Sep;60(3):296–300.

[38] Zhang MY, Wang J, Guo J. Role of Regenerating Islet-Derived Protein 3A in Gastrointestinal Cancer. Front Oncol. 2019;9:1449.

[39] Peterson KM, Buss J, Easley R, et al. REG1B as a predictor of childhood stunting in Bangladesh and Peru. Am J Clin Nutr. 2013 May;97(5):1129–33.

[40] Bein A, Fadel CW, Swenor B, et al. Nutritional deficiency in an intestine-on-a-chip recapitulates injury hallmarks associated with environmental enteric dysfunction. Nat Biomed Eng. 2022 Nov;6(11):1236–1247.

[41] Rae J, Hackney J, Huang K, et al. Identification of an IL-22-Dependent Gene Signature as a Pharmacodynamic Biomarker. International Journal of Molecular Sciences. 2021;22(15):8205.

[42] Mihi B, Gong Q, Nolan LS, et al. Interleukin-22 signaling attenuates necrotizing enterocolitis by promoting epithelial cell regeneration. Cell Rep Med. 2021 Jun 15;2(6):100320.

[43] Huang X, Zhou Y, Sun Y, et al. Intestinal fatty acid binding protein: A rising therapeutic target in lipid metabolism. Prog Lipid Res. 2022 Jul;87:101178.

[44] Lau E, Marques C, Pestana D, et al. The role of I-FABP as a biomarker of intestinal barrier dysfunction driven by gut microbiota changes in obesity. Nutrition & Metabolism. 2016;13.

[45] Reisinger KW, Derikx JPM, Thuijls G, et al. Noninvasive measurement of intestinal epithelial damage at time of refeeding can predict clinical outcome after necrotizing enterocolitis. Pediatric Research. 2013 2013/02/01;73(2):209–213.

[46] Ganz T. Defensins: antimicrobial peptides of innate immunity. Nat Rev Immunol. 2003 Sep;3(9):710–20.

[47] Spencer JD, Hains DS, Porter E, et al. Human alpha defensin 5 expression in the human kidney and urinary tract. PLoS One. 2012;7(2):e31712.

[48] Gagne D, Shajari E, Thibault MP, et al. Proteomics Profiling of Stool Samples from Preterm Neonates with SWATH/DIA Mass Spectrometry for Predicting Necrotizing Enterocolitis. Int J Mol Sci. 2022 Oct 1;23(19).

[49] Tremblay É, Thibault M-P, Ferretti E, et al. Gene expression profiling in necrotizing enterocolitis reveals pathways common to those reported in Crohn’s disease. BMC Medical Genomics. 2016 2016/01/22;9(1):6.

[50] Klerk DH, Plösch T, Verkaik-Schakel RN, et al. DNA Methylation of TLR4, VEGFA, and DEFA5 Is Associated With Necrotizing Enterocolitis in Preterm Infants. Front Pediatr. 2021;9:630817.

[51] Jin L, Yu B, Armando I, et al. Mitochondrial DNA-Mediated Inflammation in Acute Kidney Injury and Chronic Kidney Disease. Oxidative Medicine and Cellular Longevity. 2021 2021/06/30;2021:9985603.

[52] Tan K, Fujimoto M, Takii R, et al. Mitochondrial SSBP1 protects cells from proteotoxic stresses by potentiating stress-induced HSF1 transcriptional activity. Nat Commun. 2015 Mar 12;6:6580.

[53] Ferat-Osorio E, Sánchez-Anaya A, Gutiérrez-Mendoza M, et al. Heat shock protein 70 down-regulates the production of toll-like receptor-induced pro-inflammatory cytokines by a heat shock factor-1/constitutive heat shock element-binding factor-dependent mechanism. Journal of Inflammation. 2014 2014/07/12;11(1):19.

[54] Afrazi A, Sodhi CP, Good M, et al. Intracellular heat shock protein-70 negatively regulates TLR4 signaling in the newborn intestinal epithelium. J Immunol. 2012 May 1;188(9):4543–57.

[55] Ross D, Siegel D. Functions of NQO1 in Cellular Protection and CoQ10 Metabolism and its Potential Role as a Redox Sensitive Molecular Switch [Review]. Frontiers in Physiology. 2017 2017-August-24;8.

[56] Zhang Y, Wang O, Mi H, et al. Rhus chinensis Mill. fruits prevent necrotizing enterocolitis in rat pups via regulating the expressions of key proteins involved in multiple signaling pathways. J Ethnopharmacol. 2022 May 23;290:115103.

[57] Wohlschlegel J, Argentini M, Michiels C, et al. First identification of ITM2B interactome in the human retina. Scientific Reports. 2021 2021/08/26;11(1):17210.

[58] Yuan Z-C, Xu W-D, Liu X-Y, et al. Biology of IL-36 Signaling and Its Role in Systemic Inflammatory Diseases [Review]. Frontiers in Immunology. 2019 2019-October-31;10.

[59] Carrier Y, Ma H-L, Ramon HE, et al. Inter-Regulation of Th17 Cytokines and the IL-36 Cytokines In Vitro and In Vivo: Implications in Psoriasis Pathogenesis. Journal of Investigative Dermatology. 2011 2011/12/01/;131(12):2428–2437.

[60] Cho SX, Rudloff I, Lao JC, et al. Characterization of the pathoimmunology of necrotizing enterocolitis reveals novel therapeutic opportunities. Nat Commun. 2020 Nov 13;11(1):5794.

[61] Sekikawa A, Fukui H, Suzuki K, et al. Involvement of the IL-22/REG Ialpha axis in ulcerative colitis. Lab Invest. 2010 Mar;90(3):496–505.

[62] He GW, Lin L, DeMartino J, et al. Optimized human intestinal organoid model reveals interleukin-22-dependency of paneth cell formation. Cell Stem Cell. 2022 Sep 1;29(9):1333–1345.e6.

[63] Nguyen PM, Putoczki TL, Ernst M. STAT3-Activating Cytokines: A Therapeutic Opportunity for Inflammatory Bowel Disease? J Interferon Cytokine Res. 2015 May;35(5):340–50.

[64] Ma Q, Luan J, Bai Y, et al. Interleukin-22 in Renal Protection and Its Pathological Role in Kidney Diseases. Front Immunol. 2022;13:851818.

[65] Rushworth SA, MacEwan DJ, O’Connell MA. Lipopolysaccharide-induced expression of NAD(P)H:quinone oxidoreductase 1 and heme oxygenase-1 protects against excessive inflammatory responses in human monocytes. J Immunol. 2008 Nov 15;181(10):6730–7.

[66] Esmon CT. The impact of the inflammatory response on coagulation. Thrombosis Research. 2004 2004/01/01/;114(5):321–327.

[67] Mao H, Han B, Li H, et al. FABP4 knockdown suppresses inflammation, apoptosis and extracellular matrix degradation in IL-1β-induced chondrocytes by activating PPARγ to regulate the NF-κB signaling pathway. Mol Med Rep. 2021 2021/12/01;24(6):855.

[68] de Leeuw E, Rajabi M, Zou G, et al. Selective arginines are important for the antibacterial activity and host cell interaction of human alpha-defensin 5. FEBS Lett. 2009 Aug 6;583(15):2507–12.

[69] Schurink M, Scholten IG, Kooi EM, et al. Intestinal fatty acid-binding protein in neonates with imminent necrotizing enterocolitis. Neonatology. 2014;106(1):49–54.

[70] Coufal S, Kokesova A, Tlaskalova-Hogenova H, et al. Urinary I-FABP, L-FABP, TFF-3, and SAA Can Diagnose and Predict the Disease Course in Necrotizing Enterocolitis at the Early Stage of Disease. Journal of Immunology Research. 2020 2020/03/03;2020:3074313.

[71] Chen CC, Llado V, Eurich K, et al. Carbohydrate-binding motif in chitinase 3-like 1 (CHI3L1/YKL-40) specifically activates Akt signaling pathway in colonic epithelial cells. Clin Immunol. 2011 Sep;140(3):268–75.

[72] Schlapbach LJ, Aebi C, Fisch U, et al. Higher Cord Blood Levels of Mannose-Binding Lectin-Associated Serine Protease-2 in Infants With Necrotising Enterocolitis. Pediatric Research. 2008 2008/11/01;64(5):562–566.

[73] Sylvester KG, Ling XB, Liu GY, et al. Urine protein biomarkers for the diagnosis and prognosis of necrotizing enterocolitis in infants. J Pediatr. 2014 Mar;164(3):607–12 e1-7.

[74] Visscher MO, Carr AN, Narendran V. Epidermal Immunity and Function: Origin in Neonatal Skin [Review]. Frontiers in Molecular Biosciences. 2022 2022-June-08;9.

[75] Chau S, Gao J, Diao AJ, et al. Diverse yeast antiviral systems prevent lethal pathogenesis caused by the L-A mycovirus. Proceedings of the National Academy of Sciences. 2023;120(11):e2208695120.

[76] Fehr AR, Singh SA, Kerr CM, et al. The impact of PARPs and ADP-ribosylation on inflammation and host-pathogen interactions. Genes Dev. 2020 Mar 1;34(5-6):341–359.

[77] Williams D, Mahmoud M, Liu R, et al. Stable flow-induced expression of KLK10 inhibits endothelial inflammation and atherosclerosis. eLife. 2022 2022/01/11;11:e72579.

[78] Czyzyk J, Henegariu O, Preston-Hurlburt P, et al. Enhanced Anti-Serpin Antibody Activity Inhibits Autoimmune Inflammation in Type 1 Diabetes. The Journal of Immunology. 2012;188(12):6319–6327.

[79] Chamberlain PP, Qian X, Stiles AR, et al. Integration of inositol phosphate signaling pathways via human ITPK1. J Biol Chem. 2007 Sep 21;282(38):28117–25.

[80] Luke CJ, Pak SC, Askew YS, et al. An intracellular serpin regulates necrosis by inhibiting the induction and sequelae of lysosomal injury. Cell. 2007 Sep 21;130(6):1108–19.

[81] Lee YJ, Park SY, Park EK, et al. Unique cartilage matrix-associated protein regulates fibrillin-2 expression and directly interacts with fibrillin-2 protein independent of calcium binding. Biochem Biophys Res Commun. 2019 Apr 2;511(2):221–227.

[82] Lee SR, Heo JH, Jo SL, et al. Progesterone receptor membrane component 1 reduces cardiac steatosis and lipotoxicity via activation of fatty acid oxidation and mitochondrial respiration. Sci Rep. 2021 Apr 22;11(1):8781.

[83] Luo K, Zhang L, Liao Y, et al. Effects and mechanisms of Eps8 on the biological behaviour of malignant tumours (Review). Oncol Rep. 2021 2021/03/01;45(3):824–834.

[84] Pan W, Nagpal K, Suárez-Fueyo A, et al. The Regulatory Subunit PPP2R2A of PP2A Enhances Th1 and Th17 Differentiation through Activation of the GEF-H1/RhoA/ROCK Signaling Pathway. J Immunol. 2021 Apr 15;206(8):1719–1728.

[85] Kerr SC, Fieger CB, Snapp KR, et al. Endoglycan, a member of the CD34 family of sialomucins, is a ligand for the vascular selectins. J Immunol. 2008 Jul 15;181(2):1480–90.

[86] Zhu C, Liu C, Chai Z. Role of the PADI family in inflammatory autoimmune diseases and cancers: A systematic review. Front Immunol. 2023;14:1115794.

[87] Abbasoglu A, Sarialioglu F, Yazici N, et al. Serum Neuron-specific Enolase Levels in Preterm and Term Newborns and in Infants 1–3 Months of Age. Pediatrics & Neonatology. 2015 2015/04/01/;56(2):114–119.

[88] Lin SN, Musso A, Wang J, et al. Human intestinal myofibroblasts deposited collagen VI enhances adhesiveness for T cells - A novel mechanism for maintenance of intestinal inflammation. Matrix Biol. 2022 Nov;113:1–21.

[89] Lin H, Zeng W, Lei Y, et al. Tuftelin 1 (TUFT1) Promotes the Proliferation and Migration of Renal Cell Carcinoma via PI3K/AKT Signaling Pathway. Pathol Oncol Res. 2021;27:640936.

[90] Grover R, Burse SA, Shankrit S, et al. Myg1 exonuclease couples the nuclear and mitochondrial translational programs through RNA processing. Nucleic Acids Res. 2019 Jun 20;47(11):5852–5866.

[91] Ma J, Wei K, Liu J, et al. Glycogen metabolism regulates macrophage-mediated acute inflammatory responses. Nat Commun. 2020 Apr 14;11(1):1769.

[92] Koyama S, Akbay EA, Li YY, et al. STK11/LKB1 Deficiency Promotes Neutrophil Recruitment and Proinflammatory Cytokine Production to Suppress T-cell Activity in the Lung Tumor Microenvironment. Cancer Res. 2016 Mar 1;76(5):999–1008.

[93] Sodhi CP, Shi XH, Richardson WM, et al. Toll-like receptor-4 inhibits enterocyte proliferation via impaired beta-catenin signaling in necrotizing enterocolitis. Gastroenterology. 2010 Jan;138(1):185–96.

